# Development and validation of a quantitative instrument for measuring temporal and social disorientation in the Covid-19 crisis

**DOI:** 10.1101/2022.02.28.22270969

**Authors:** Pablo Fernandez Velasco, Umer Gurchani, Bastien Perroy, Tom Pelletreau-Duris, Roberto Casati

## Abstract

We developed a quantitative Instrument for measuring Temporal and Social Disorientation (ITSD), aimed at major crises such as the Covid-19 pandemic. Disorientation has been identified as one of the central elements of the psychological impact of the Covid-19 era on the general public, but so far, the question has only been approached qualitatively. This paper offers an empirical, quantitative approach to the multi-faceted disorientation of the Covid-19 pandemic by operationalising the issue with the help of the ITSD. The ITSD was developed through multiple stages involving a preliminary open-ended questionnaire followed by a coder-based thematic analysis. This paper establishes the reliability and validity of the resulting ITSD using a 3-step validation process on a sample size of 3306.

## Introduction

The Covid-19 crisis saw the breakdown of many of the landmarks we commonly use to orient ourselves in our daily life. Social, spatial, and temporal disruptions, such as curfews, social distancing measures and repeated lockdowns (both current and potential), suddenly supersede the previous organization of life in sometimes confusing ways. Many authors have claimed that disorientation is a central element in people’s experience during the Covid-19 era [1, 2, 3, 4], with contributions coming from social psychology [5], geography [6], philosophy [7], ethnography [8] or cultural studies [9], to name but a few disciplines. Such discussions are not only in the purview of academic research, but are also common in mainstream culture, with the media explicitly discussing the disorienting nature of lockdowns [10], of reopening [11], of routine disruption [12], of Covid-19 data [13], of new waves [14], and of the pandemic as a whole [15].

So far, the resulting disorientation of the Covid-19 pandemic has been addressed qualitatively through various conceptual, ethnographic, or phenomenological approaches. The aim of this paper is to operationalise this research question by developing and validating an instrument that can test empirically and quantitively the multi-faceted disorientation of the Covid-19 pandemic.

From a preliminary theoretical standpoint, if spatial disorientation is the metacognitive feeling that emerges when one has difficulty locating oneself within a spatial frame of reference [16], then, accordingly, temporal and social disorientation during Covid would be such a feeling relative to organizing one’s time or knowing one’s place within temporal and social frames of reference [3]. In order to assess the extent to which the Covid-19 crisis was a disorienting period, we developed a tool to measure the experienced cognitive distortions related to the self-locational aspects of our cognition within these various frames of reference: the Instrument for Measuring Temporal and Social Disorientation (ITSD).

In what follows, we will first describe the development of the ITSD, then assess its empirical validation on a large sample size (n = 3306), and finally discuss the main cognitive components that emerged through the validation process — a unified social disorientation construct, plus a range of separate aspects of temporal disorientation. Although the ITSD has been developed for the Covid 19 crisis, the instrument can be adapted for quantifying the disorienting impact of any future crisis.

## Materials and Methods

We developed the quantitative instrument in three stages and then validated it using a further three steps. First, we created an open-ended, qualitative questionnaire. Second, we performed a thematic analysis of the results of said qualitative questionnaire. Third, we developed a first draft of the quantitative instrument for which we received feedback from four experts in different relevant fields. We adapted the final version of the quantitative instrument based on their recommendations before sharing the questionnaire online through an emailing campaign as well as through Twitter.

With the initial open-ended qualitative questionnaire, we wanted to probe the variety of experiences relating to temporal, social and epistemic disorientation. At the same time, we wanted to phrase our questions in a sufficiently indirect way to avoid priming subjects. As a result, we included three separate, open-ended questions asking participants to outline any time distortions they might have felt during the pandemic, any way in which they might have felt socially out of place, and any way in which they might have felt at a loss in navigating the information related to the Covid-19 crisis, respectively. Each of these questions included several examples to help the participants understand what was being asked of them. After distributing the questionnaire through emails in universities mailing lists in France and in the UK in the course of March 2021, 161 participants completed at least one of these three open-ended questions (98 of whom were French, 63 of whom were English).

We performed different levels of thematic analysis for the responses of each of the three questions. We found that the responses for social disorientation were coherent and homogeneous, and we easily agreed on the emerging 9 Likert-scale questions for the quantitative instrument, which we hypothesized would all form a single component. In contrast, the responses for the question related to epistemic disorientation were very heterogeneous, and we decided to exclude the construct from the quantitative instrument.

Temporal disruptions required the most detailed analysis. We found that a variety of temporal disruptions were consistently reported in the open questionnaire, so we decided to undertake further, more fine-grained analysis using two independent coders and following some established thematic analysis guidelines [17, 18]. First, each of the coders tagged all the reports with descriptions of the experience reported by each respondent. Then, the coders compared tags and agreed on a list of tags, organized in a taxonomical tree spanning 5 overarching categories (e.g., time distortion), 9 subcategories (e.g., passage of time) and 40 individual tags (e.g., time passing slower).

We divided the participant’s responses into units of meaning by having the coder most specialized in temporal disorientation retrieve all of his spontaneous tags, clear his tags while keeping the boundaries of what had been tagged, and add subsequent divisions for the sections of the participant’s responses that were untagged. The corpus was composed at this stage of 358 units of meaning across 149 participants having answered the temporal question (95 of whom were French, 54 of whom were English). The questionnaire was fully anonymous, and participants completed a written consent form online. All research involving human participants has followed the ethical procedures for approval at our institution. The Pôle Éthique of the Institut des Sciences Biologiques (INSB) of the Centre national de la recherche scientifique (CNRS) waived all ethical approval for fully anonymous questionnaires. The study has been conducted according to the principles expressed in the Declaration of Helsinki.

The two coders then blindly tagged each unit of meaning by ticking yes/no for each of the 40 individual tags. This resulted in 96.30% agreement across coders (including null agreements). Excluding null agreements (i.e. which makes agreement happen if and only if both coders tagged a unit with the same tag, not if they agree not to tag an unit with a given tag, as following Miles and Huberman 1984 intercoder agreement metric), resulted in 50.92% agreement across coders. Such a rate is standard for complex coding tables due to the cognitive difficulty to monitor that many tags for that many units of meaning at once [17]. This is why we undertook a second round of re-tagging (in which the coders could assess the other coder’s tags, and blindly change one’s own tags), which resulted in 90.28% agreement rate (excluding null agreements). Note that re-assessing each tag blindly might have worsened the agreement rate in some cases, e.g. if both coders changed their opinion at once on a given disagreement.

The relatively high level of agreement at each of these steps of the thematic analysis led us to consider the tagged states to be good candidates for the quantitative instruments, and we created 24 questions based on them, which we hypothesized would correspond to six separate components, given the heterogeneity of temporal disruptions. Of the 40 tags in the coding process, 10 had to do with temporal scale (e.g. day-scale vs month-scale) and not with particular experiences of temporal distortions, so they did not become questions in the quantitative instrument. Five other tags had to do with psychological disruptions that were not strictly temporal and that were better addressed through the Global Psychotrauma Screen (e.g. anxiety) or through demographic and lifestyle questions (e.g. routine disruption) and were also discarded, which resulted in the final selection of 24 questions related to temporal disruptions.

We then added questions about demographic and lifestyle changes, as well as the full Global Psychotrauma Screen (GPS) and the MacArthur Scale of Subjective Social Status. We decided to use GPS because it had been previously validated and because it already incorporated many elements that had emerged in the responses to the open-ended questionnaire, such as anxiety, boredom, or derealisation. Moreover, it had previously been adapted to the context of the Covid-19 pandemic [19].

We proceeded to send the quantitative instrument to four separate experts for feedback: two psychologists, a human geographer specialising on disorientation, and a philosopher in the phenomenological tradition. We rephrased some questions based on their feedback, and we eliminated 12 questions (e.g., the last four questions in the GPS), partly because the experts suggested that a large number of questions might hamper participation.

The final version of the quantitative instrument included 9 demographic questions, the MacArthur Scale of Subjective Social Status, 11 questions on lifestyle changes (pre- vs post-pandemic), 9 social disorientation questions, 24 temporal disorientation questions (divided into three blocks) and 13 questions from the GPS. The questionnaire was hosted on Qualtrics.

When it came to distributing the questionnaire, we decided to only focus on online distribution, as the restrictions meant that physical distribution was ill-suited for our purposes. Higher-education students have been described as an at-risk population from a psychological standpoint during the Covid-19 crisis [20]. And as reports from students were among the most disrupted ones we noticed in the qualitative questionnaire, we decided to move further with a strategy geared towards this population. We sent an email to 105 communication directors of major higher education institutions in France to ask them to share the questionnaire with their students. In order to diversify our sample, we also distributed the questionnaire on Twitter. Having two different distribution strategies aimed at different groups also increases our ability to contrast responses from different demographics. We shared a link to the questionnaire to active French users (i.e. tweeting about anything) on the social network, inviting them to share the questionnaire with their own followers.

The questionnaire was distributed through both email (4724 respondents) and Twitter (727 respondents) in May and June 2021, just after the third national lockdown ended, at a time of an ongoing 6-month-old curfew that was preceded by the 7 weeks of the second national lockdown in late 2020. (During the first semester of 2021 in France, lockdown and curfew measures were combined.) Hence, at the time they were queried, our respondents have gone through around 7 months of heavy social, spatial and temporal restrictions. There was no financial compensation for completing the questionnaire. The sample size was 5453 participants, including those who only partially answered the survey. 3306 participants completed the full survey. The questionnaire was fully anonymous, and participants completed a written consent form online. All research involving human participants has followed the ethical procedures for approval at our institution and has been conducted according to the principles expressed in the Declaration of Helsinki. As a result of our distribution strategy, most of our sample is composed of higher-education students despite the marginal presence of workers or retirees, which is captured by a median age of 21 and an average age of 25 in our validation sample.

In order to establish reliability and construct validity of the ITSD we followed ‘*the 3 faced construct validation method*’ using Factor Analysis [21]. We divided the sample of people having answered the questionnaire completely into 3 subsamples at random (20%, 40%, 40%). In the first step we used the 20 percent sample for standard exploratory factor analysis (EFA) and calculated Cronbach’s alpha value for each of the factor detected. In the second step we used the first 40 percent subsample to perform CFA to find the most optimum model, and the third step was cross-validation of the optimal model with another CFA on the last 40 percent subsample to avoid over-fitting issues.

## Results

We began the validation process of the ITSD by running an exploratory Factor Analysis on the first 20 percent subsample, which was a first step towards understanding the underlying components of the data. For running EFA we only took questions related to social-disorientation and temporal disorientation. In order to ensure the suitability of data for Factor Analysis, the following two tests were carried out:

1. Kaiser-Meyer-Olkin
2. Bartlett’s test of sphericity

Bartlett’s test of sphericity [22] tests the overall significance of all the correlations within the correlation matrix, was found to be significant (χ 2 (3306) = 5187.95, p < 0.0001) which made it clear that the data was appropriate for using Factor Analysis. The Kaiser-Meyer-Olkin measure of sampling adequacy indicated that the strength of the relationships among variables was also high enough to run Factor analysis (KMO = 0.830). Once these prerequisites were met, we used ‘FactorAnalyzer’ module in Python to run the Factor Analysis. An oblique rotation was chosen for factor analysis as we anticipated that the components may be correlated. Figure 1 shows the scree plot and eigen-values for corresponding components discovered.

**Figure 1:**
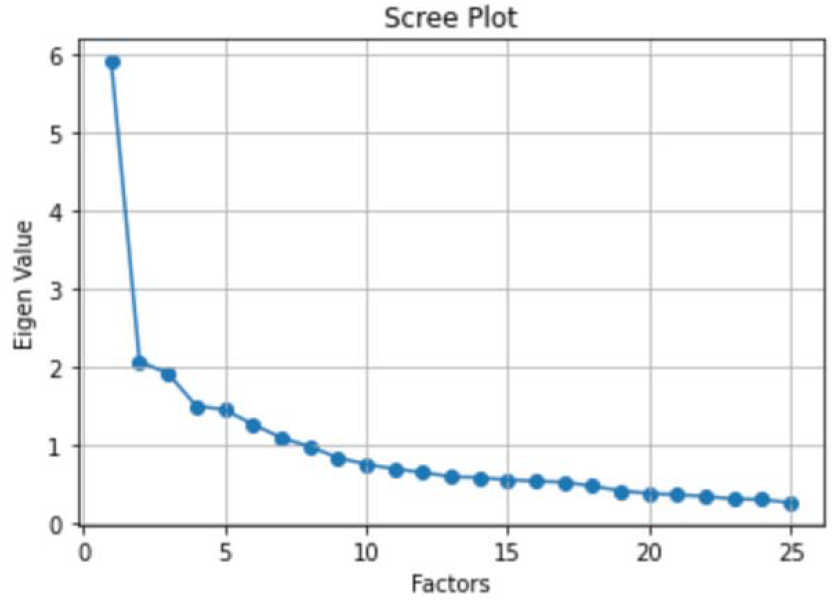
Scree-Plot for the EFA

**Figure 2:**
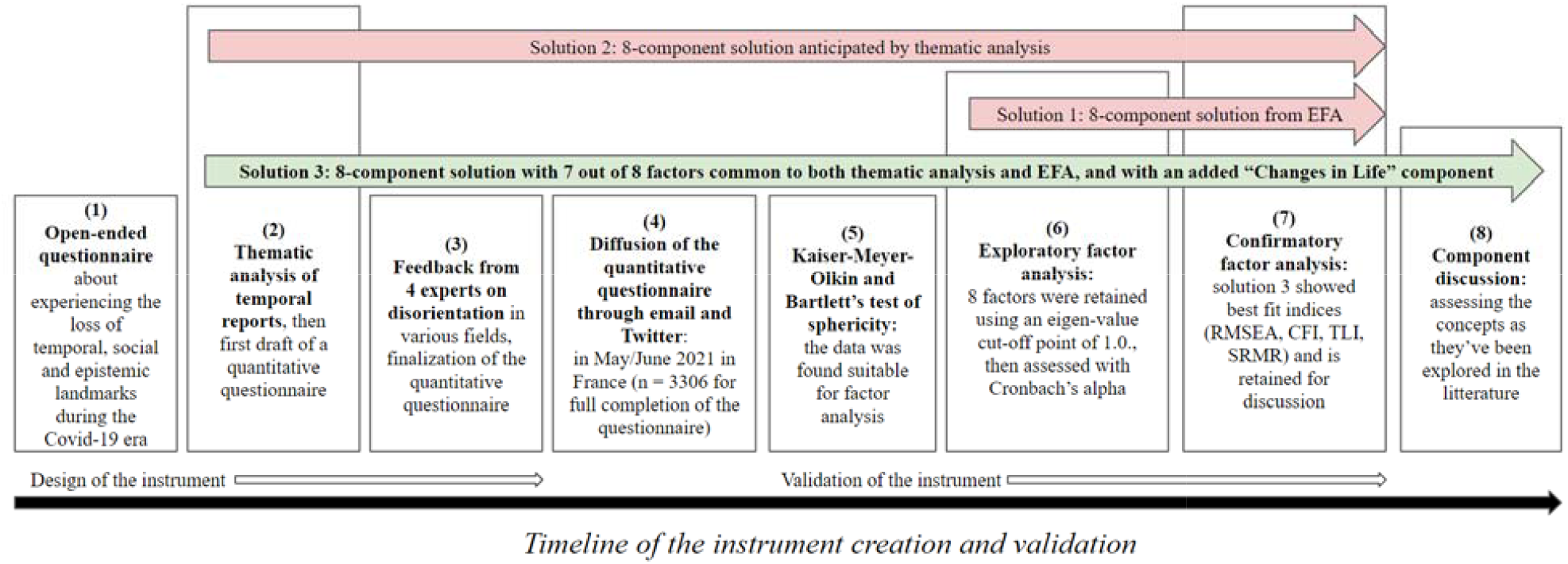
flow-chart of the instrument design and validation process

From the EFA result we took the first 8 components as they explained more than 46 percent of the variance in the date. Cronbach’s α was calculated to evaluate internal consistency of the components found through Factor analysis. An α-level of 0.05 was used to determine statistical significance. Eigenvalues for each component are (5.9, 2.07, 1.92, 1.5, 1.46, 1.27, 1.1, 0.99). Component loadings and item statistics are presented in *Table 4*, in the order in which they were presented to respondents. As mentioned above, temporal questions were divided into three sections, two of them being associated with a temporal framing. People had 5 Likert-scale possible answers, e.g. “I feel I’m [way more / more / neither more nor less / less / way less] reliant on calendars or to-do lists to keep track of what I do”. Items for which no underscored adverbs are outlined were asked with a [strongly agree / agree / neutral / disagree / strongly disagree] set of possible answers. As a result of EFA (without any influence from thematic analysis), we identified the components as following after careful examination:

1. SD: Social Disorientation (Questions 20, 21, 22, 23, 24, 27)
2. PT: Passage of Time (Questions 48, 49, 50)
3. TOE: Temporal Order of Events (Questions 42, 43, 54)
4. TD: Temporal Distance (Questions 45, 46, 47)
5. FO: Future Orientation (Questions 52, 56, 57)
6. TSL: Temporal Self Location (Questions 40, 41)
7. ATO: Assisted Temporal Orientation (Questions 36, 37)
8. TR: Temporal Rupture (Questions 44, 58)

**Table 1:** Sample Characteristics of the 3306 participants having answered all questions

| *Characteristics* | *Values* |
| --- | --- |
| *Average Age* | 25.24 |
| *Median age* | 21.00 |
| *Number of Female Participants* | 2273 |
| *Number of Male Participants* | 990 |
| *Number of Participants that chose neither*<br>*‘male’ nor ‘female’* | 43 |
| *Average Socio-Economic Status (based on*<br>*the MacArthur Scale of Subjective Social*<br>*Status)* | 6.04 |
| *Number of students* | 2392 |
| *Number of workers* | 342 |
| *Number of retirees* | 64 |
| *Number of unemployed* | 25 |
| *Number of ‘chômage partiel’ (a French*<br>*special status during the crisis, with*<br>*wages mostly covered by the state and*<br>*the person not actively working)* | 3 |
| *Average Number of Cohabitants* | 2.3 |

**Table 2:** Factor Loadings for Temporal Disorientation Components. Questions were asked in French (see the full original questionnaire in the supplementary materials); underlined are adverbs used as a Likert-scale set of 5 answers; boldened factor loadings indicate questions used for the component in both solution 2 and solution 3.

| Items (questions) | PT | TOE | TD | FO | TSL | ATO | TR |
| --- | --- | --- | --- | --- | --- | --- | --- |
| **Section I** |  |  |  |  |  |  |  |
| Since the beginning of the Covid-19 crisis... |  |  |  |  |  |  |  |
| Q36. I feel I’m more / less reliant on calendars or to-do lists to keep track of what I do. | -0.002 | -0.050 | -0.076 | 0.006 | 0.048 | 0.992 | -0.054 |
| Q37. I care more / less about following a routine (daily, or weekly). | 0.033 | 0.019 | 0.017 | 0.050 | -0.108 | 0.397 | 0.011 |
| Q38. I feel that more / less activities or tasks would<br>be doable in a day or in a week. | 0.028 | -0.007 | 0.021 | 0.239 | -0.255 | 0.149 | 0.047 |
| Q39. I feel more / less late overall on my<br>commitments or my deadlines. | -0.047 | -0.052 | -0.040 | -0.199 | 0.276 | -0.077 | -0.037 |
| Q40. I get confused more / less often about which<br>day of the week it is. | -0.059 | -0.202 | -0.067 | -0.082 | 0.431 | -0.012 | -0.078 |
| Q41. I get confused more / less often about which<br>month of the year it is. | 0.001 | -0.141 | -0.032 | -0.033 | 0.985 | 0.038 | -0.079 |
| Section II |  |  |  |  |  |  |  |
| Q42. At times, I feel confused about the order of<br>events that occurred since the pandemic began. | 0.001 | 0.68 | 0.060 | 0.040 | -0.256 | -0.027 | 0.152 |
| Q43. At times, I feel confused about the order of<br>events that occurred before the pandemic began. | 0.006 | 0.651 | 0.033 | -0.027 | -0.045 | -0.035 | 0.048 |
| Q44. The period since the pandemic began feel<br>connected / disconnected from the months and years<br>prior. | 0.047 | 0.072 | 0.155 | 0.139 | -0.112 | -0.042 | 0.44 |
| Q45. At times, the beginning of the pandemic feels<br>noticeably far away. | 0.036 | 0.194 | 0.674 | -0.012 | -0.106 | -0.075 | 0.180 |
| Q46. At times, the beginning of the pandemic feels<br>noticeably close. | -0.124 | 0.031 | 0.676 | -0.082 | -0.030 | 0.015 | 0.091 |
| Q47. Overall, times before the pandemic feel as if<br>they are further away / closer to me than they really<br>are. | 0.086 | 0.169 | 0.547 | 0.081 | -0.090 | -0.047 | 0.198 |
| Q48. At times, since the pandemic began, time has<br>been passing noticeably slowly. | 0.728 | 0.067 | 0.087 | 0.000 | -0.118 | -0.028 | 0.151 |
| Q49. At times, since the pandemic began, time has<br>been passing noticeably quickly. | 0.697 | -0.060 | 0.081 | 0.091 | 0.063 | 0.056 | -0.077 |
| Q50. Overall, since the pandemic began, time has<br>been passing slowly / quickly. | 0.828 | 0.021 | 0.074 | 0.028 | -0.052 | -0.007 | 0.042 |
| Section III |  |  |  |  |  |  |  |
| Since the beginning of the Covid-19 crisis... |  |  |  |  |  |  |  |
| Q52. I feel it is easier / harder to imagine for me to<br>imagine the future. | 0.054 | 0.119 | 0.074 | 0.720 | -0.063 | 0.020 | 0.148 |
| Q53. I feel it is easier / harder to imagine for me to recall events having taken place since the pandemic began. | -0.008 | 0.317 | 0.101 | 0.164 | -0.214 | -0.001 | 0.066 |
| Q54. I feel it is easier / harder to imagine for me to recall events having taken place before the pandemic began. | 0.003 | 0.397 | 0.138 | 0.126 | -0.072 | -0.021 | 0.028 |
| Q55. I feel I ruminate more / less about past events emotionally negatively charged. | 0.094 | 0.068 | 0.084 | **0.327** | -0.106 | -0.015 | 0.105 |
| Q56. I feel I'm more / less anxious about my future. | 0.060 | 0.084 | 0.092 | **0.640** | -0.060 | -0.029 | 0.140 |
| Q57. I feel I'm more / less in control of my future. | 0.059 | 0.102 | 0.060 | **0.565** | -0.100 | 0.036 | 0.041 |
| Q58. At times, the period since the pandemic began felt unreal to me. | 0.043 | 0.139 | 0.094 | 0.084 | -0.101 | -0.059 | **0.490** |

**Table 3:**
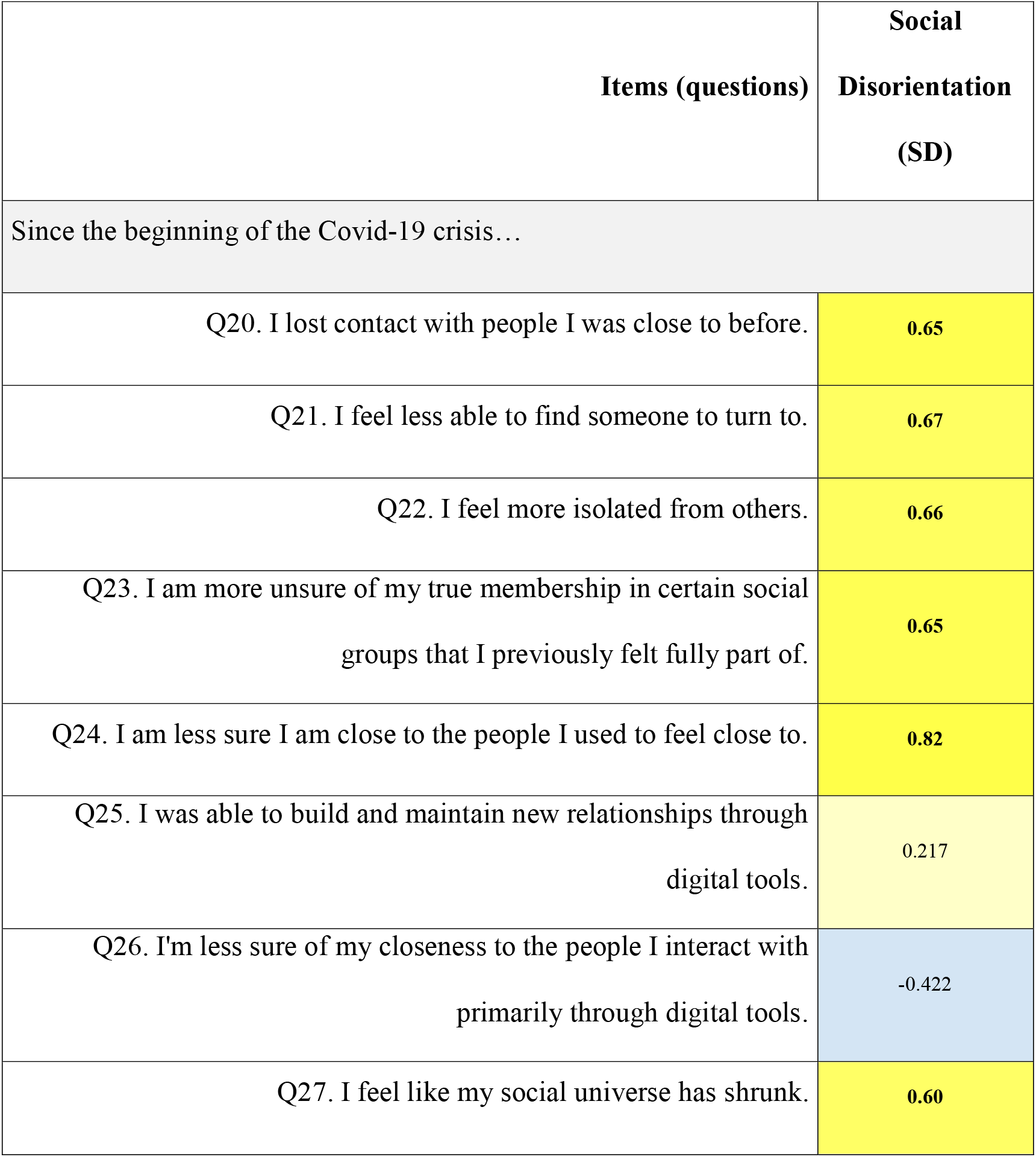
Factor Loadings for Social Disorientation Component. Questions were asked in French (see the full original questionnaire in the supplementary materials); questions were asked with a Likert-scale set of 5 possible answers; boldened factor loadings indicate questions used for the SD in both solution 2 and solution 3.

**Table 4:** Cronbach’s Alpha Values for components

| Construct | Cronbach’s<br>alpha value |
| --- | --- |
| Social Disorientation Factor |  |
| Social Disorientation (SD) | 0.865 |
| Temporal Disorientation Factors |  |
| Passage of time (PT) | 0.80 |
| Temporal Order of Events (TOE) | 0.63 |
| Temporal Distance (TD) | 0.65 |
| Future Orientation (FO) | 0.74 |
| Temporal Self Location (TSL) | 0.65 |
| Assisted Temporal Orientation (ATO) | 0.52 |
| Temporal Rupture (TR) | 0.41 |

**Table 5:** Fit indices for solution 3

| RMSEA | CFI | TLI | SRMR |
| --- | --- | --- | --- |
| 0.052 | 0.90 | 0.89 | 0.048 |

### Reliability

To establish the internal consistency, we calculated Cronbach’s alpha values for each of the components detected in EFA. Table 4 shows the result of these tests.

### Confirmatory Factor Analysis

As we had anticipated a component structure through thematic analysis and there were some differences between it and EFA results, we performed a confirmatory factor analysis to find the best possible component structure that resolved the differences between the two. The first Confirmatory factor analysis (40% subsample) was performed on an 8-component solution proposed by EFA (solution 1), and 8 component solutions anticipated by thematic analysis of the qualitative questionnaire (solution 2). The only difference between these two solutions was the composition of factor ‘Future Orientation’. In the first solution we did not include the question, “I feel I ruminate more / less about past events” in ‘future orientation’ as the factor loading value for it was very low compared to other questions included in this latent variable. However, in the second solution we included this question in ‘future orientation’ as anticipated in our thematic analysis. Neither of the above two solutions presented a good fit. Consequently, a third solution with 8 factors was also tested, which included common factors from both thematic analysis and EFA, except for one factor being removed and another one being added in its place. ‘Lifestyle Changes’ factor was added, which was constructed by subtracting the level of physical activity and time spent at home during Covid-19 from pre-Covid physical activity and time spent at home. ‘Assisted temporal orientation’, found in both EFA (but for which we had a relatively low Cronbach’s alpha value) and thematic analysis (but for which the associated tag wasn’t a very prominent tag), was also removed due to CFA fit indices turning out to be better without it. Despite its low Cronbach alpha value, we decided to keep ‘Temporal rupture’ in solution 3 due to its distinctiveness observed in thematic analysis. Like solution 2, in solution 3 we included “I feel I ruminate more / less about past events” in ‘future orientation’. For solution 3 we found the following fit indices.

The above results were then replicated on the last subsample of 40 percent which confirmed that solution 3 which is a mixture of EFA and thematic analysis (and ignores Assisted Temporal Orientation) showed better fit indices (*RMSEA < 0*.*05, CFI>0*.*88, TLI>0*.*88, SRMR<0*.*05)*.

## Discussion

From the validation process described in the previous section, solution 3 showed the best fit indices. In this solution, we found 8 components, which we are now going to describe from a theoretical point of view. The final component structure includes social disorientation as a single component, six temporal components underpinning temporal disorientation, as well as a “lifestyle changes” component that we generated from a comparison of lifestyle questions symmetrically asked for before and after the beginning of the crisis.

Most of the temporal components we found are congruent with the thematic analysis. In the qualitative reports, *distortions of the feeling of passage of time, difficulties to project oneself into the future, difficulties to locate oneself in time*, and *distortions of subjective temporal distances* were respectively the four most often reported phenomena. These all made it into strong components within the above-mentioned validation process of the quantitative data. *Temporal rupture* was a somewhat rarer phenomenon in the qualitative report, but its distinctiveness led us to believe it would probably turn out to be a noteworthy component, which it did.

Two dissimilarities emerge from the picture drawn from thematic analytic. On the one hand, there was one component that, based on the qualitative reports, we had not foreseen would emerge the way it did: *confusion about the temporal order of events* was a very rare phenomenon in the qualitative reports, which led us to believe that it would not make it into a strong temporal component, but it eventually did. This component almost ended up encapsulating questions about difficulties to recall past events, which were more frequent in the qualitative reports. These questions ultimately didn’t make their way into any component. On the other hand, two questions about one’s agentive feelings were minor but noteworthy phenomena spotted in thematic analysis yet didn’t make their way into any component. A question about feelings of lessened productivity almost ended up being encapsulated in *future orientation*. A somewhat related question about feelings of lateness with regards to one’s commitments almost made its way into *temporal self location*. Importantly, no major frequent phenomenon from the qualitative questionnaire, as assessed by thematic analysis, did not make its way into a temporal component – notwithstanding some phenomena that we ultimately probed through the Global Psycho Trauma section of the questionnaire rather than through our own newly created sections (for example and most importantly: monotony).

Among the two coders, there was an overall good agreement that disorientation (which also took the form of possible tags) was associated with combinations of disruptions encapsulated by the following temporal components.

Let us now proceed to provide descriptions of each component in turn.

### Social Disorientation (SD)

Social disorientation is a hindrance to the capacity to find one’s place in the social world. It involves an uncertainty regarding one’s affinity to other individuals and one’s belonging to particular social groups, as well as a sense of isolation and of one’s social world shrinking. A large international study about how Covid-19 affected felt temporalities noticeably found that social isolation captured many of the distortions of temporal experiences [23].

### Temporal Disorientation

Temporal disorientation is a feeling of uneasiness regarding the junction of perception and action in time, and with respect to one’s relative location within surrounding events. Distortions of the following 6 components can be conducive to temporal disorientation.

#### 1. Passage of time (PT)

Passage of time judgments are among the most documented aspects of one’s temporal experience. Crucially, it is now the consensus in the literature to distinguish passage of time judgments (e.g. time feeling like passing “faster” or “slower” than usual) from the production of time intervals (e.g. knowing intuitively how much time passed since a recent event) [24]. While the production of time intervals has been found to be unaffected by lockdowns [25], most Covid-19 studies report important distortions in the feeling of passage of time related to Covid-19 life constraints (e.g. both faster and slower in the UK in [26]; mainly slower in France in [27]; mainly faster in Uruguay in [28]).

#### 2. Temporal Order of events (TOE)

Time appears confusing when the temporal order of events one has in mind is incoherent. The temporal order of events is confusing when one is mistaken about which event happened before or after another, something that either violates a mental model for which causation is key (as causality is temporally asymmetric) or violates transitivity (e.g. I learn that an event x happened before an event y and after an event w, yet I believe that y precedes w). Psychologically, the ordering of events as they are remembered heavily relies on contextual inference [29, 30]. Interestingly, from our data we found this component to be composed of two questions on whether the order of events before or after the beginning of the pandemic felt confusing, as well as a question about whether events before the beginning of the pandemic were harder/easier to recall (i.e. the question about whether events after the beginning of the pandemic were harder/easier to recall did not make it into the component).

#### 3. Temporal Distance (TD)

Temporal distance is a blurrier construct within the literature than the previous two. It is closely associated with passage of time, as it refers to an intuitive feeling of the temporal distance between oneself and an event [31]. In our questionnaire, it refers to questions about whether at times the beginning of the pandemic felt close or far away, and whether overall it feels closer or farther away than what it really is. We distinguished temporal distance from passage of time from the thematic analysis onwards because some respondents of the qualitative questionnaires held contradictory statements if the two components were one and the same (e.g. time feeling elongated while also passing faster, and as it has also been reported by other researchers with statements such as ““it has already been x weeks since the beginning of the lockdown”“yet concomitant with ““it feels as if it started a year ago”“; in [32]). This discrepancy was also found by other researchers during Covid-19, noting that feeling of the passage of time for a day or a week on the one hand, and feeling of the passage of time for a year on the other most likely differ in their cognitive underpinnings, as suggested by different patterns of empirical data [33].

#### 4. Future Orientation (FO)

Future orientation is an individual’s propensity to have positive or negative future oriented mental states such as attitudes, emotions, and imaginings. It is, accordingly, a type of temporal perspective. In the literature, temporal perspectives are usually divided between the past, the present, and the future on one dimension; as well as valence (i.e. the positivity or negativity of mental states) or associated fine-grained emotional aspects on another dimension (e.g. [34]). During Covid-19, temporal perspectives have been found to be associated with different levels of compliance with restrictions [35], as well as good predictors of anxiety and depression [36], so that disorientation with respect to various temporal perspectives might actually be an important epidemiological variable worth monitoring [37]. Future orientation has also been found to be related to mental health during Covid-19, with young people being most in need of interventions designed “to restore a safe, more certain future” [38].

#### 5. Temporal Self Location (TSL)

Temporal self-location consists in knowing what hour of the day, what day of the week, or what month of the year it currently is. It has been found to be a direct source of temporal disorientation during the crisis as documented by the very first lockdown studies (e.g. [39]), in which people reported to be confused twice as much compared to before the first lockdown when they wanted to know what day of the week it was. Temporal self-location implies a continuous monitoring of the passage of time in working memory, which has been found to be affected by the Covid-19 constraints, especially time-based recalls (e.g. recalling to phone someone at a given hour of the day) [40].

#### 6. Temporal Rupture (TR)

Temporal rupture happens when one faces an unexpected far-reaching event that unsettles the whole mental timeline onto which one locates oneself [41]. In our questionnaire, it is characterized by questions about feeling a temporal disconnection before and after the outbreak of Covid-19, as well as a feeling that time has been unreal since the beginning of the pandemic.

### Depersonalization/Derealization (DP)

Depersonalization/derealization (DP/DR) is a component found within the Global Psychotrauma Screen. It designates one’s alien detachment with oneself and one’s surroundings and often comes with feelings of disembodiment, emotional numbing, or anomalous subjective recall [42]. Questions identified with DP/DR were about self-harm, life feeling dream-like or unreal, as well as feeling detached from one’s body.

### Lifestyle Changes (LC)

In order to assess lifestyle changes, three aspects were considered:

1. Changes in physical activity
2. Amount of time spent at home
3. Perception of stability in routine

‘Changes in physical activity’ and ‘amount of time spent at home’ were assessed using Likert scale questions in two categories, ‘before Covid’ and ‘after Covid’. The responses were then subtracted (‘after Covid’ – ‘before Covid’) to construct the final version of the ‘Lifestyle Changes’ component. Finally, ‘perception of stability in routine’ was measured through a direct inquiry.

Table 6 presents Pearson product-moment correlation coefficients between latent variables. Further work is needed based on our data to control important variables before putting forward any claim.

**Table 6:** Correlation Matrix for latent variables. All correlation coefficients are significant (p < 0.001).

| Latent variables | TSL | TOE | TR | TD | PT | SD | FO | LC |
| --- | --- | --- | --- | --- | --- | --- | --- | --- |
| Temporal Self Location (TSL) | --- |  |  |  |  |  |  |  |
| Temporal Order of Events (TOE) | 0.50 | --- |  |  |  |  |  |  |
| Temporal Rupture (TR) | 0.46 | 0.53 | --- |  |  |  |  |  |
| Temporal Distance (TD) | 0.30 | 0.35 | 0.54 | --- |  |  |  |  |
| Passage of Time (PT) | 0.17 | 0.11 | 0.25 | 0.26 | --- |  |  |  |
| Social Disorientation (SD) | 0.37 | 0.39 | 0.47 | 0.29 | 0.26 | --- |  |  |
| Future Orientation (FO) | -0.42 | -0.39 | -0.62 | -0.38 | -0.26 | -0.59 | --- |  |
| Lifestyle Changes (LC) | -0.21 | -0.09 | -0.19 | -0.14 | -0.10 | -0.19 | 0.18 | --- |

Confirmatory factor analysis established discriminant validity for our constructs: they capture distinctive latent variables in a reliable way (as assessed with Cronbach’s alpha). Further efforts are needed to make the components more exhaustive, as we can’t claim convergent construct validity. Our research topic being disorientation, many of our questions were designed in order for people to be able to express contradictory feelings (e.g. feeling that at times, time has been passing faster while also feeling that overall, time has been passing slower) which as we described, has been stressed by some researchers about temporal experiences associated with the pandemic. As a result, many questions are expressed in a very similar way. Hence, our questions don’t necessarily express all the possible ways for these constructs to be expressed. For instance, during thematic analysis, we found some reports about time dilation or t me compression which subsequently made us draft questions about them for ITSD. Feedback from our experts led us to believe many respondents wouldn’t grasp intuitively the distinction between compression or dilation and time feeling far or close to oneself, which led us to remove these questions. We also didn’t want to hamper participation with more questions than most people would tolerate to answer diligently. All in all, more work remains needed to establish conversion validity of these constructs, with other framings of the concepts.

## Conclusion

In this paper we validated our Instrument for measuring Social and Temporal Disorientation (ITSD) by assessing its reliability through a 3-step validation process. We did so based on a sample size of 3306 in May and June 2021 in France during a period of important Covid-19 restrictions (i.e. just after the third national lockdown, at a time of an ongoing 6 month old curfew that was preceded by the 7 weeks of the second national lockdown in late 2020). This tool was developed through the thematic analysis of reports gathered with a qualitative questionnaire about cognitive distortions in the temporal and social domains one year after the Covid-19 outbreak, in February and March 2021. From the validation data, we construed social disorientation as a hindrance to find one’s place in one’s social world; and temporal disorientation as an uneasiness about the junction of perception and action in time with respect to one’s relative location with surrounding events. Moreover, the six components related to temporal disorientation emerging from our analysis capture fine-grained distinctions within the wide range of temporal disruptions respondents might have experienced. The ITSD can be used reliably in crises that shatter the landmarks that people commonly use to orient themselves temporally and socially, such as the Covid-19 whose dimensions it seeks to characterize.

## Data Availability

The data is held in a public repository: https://www.kaggle.com/gurchani/confirmatory-factor-analysis-plos-one https://www.kaggle.com/gurchani/discovid-instrument-validation-plos-one https://www.kaggle.com/gurchani/covid19disorientationsurvey

https://www.kaggle.com/gurchani/confirmatory-factor-analysis-plos-one

https://www.kaggle.com/gurchani/discovid-instrument-validation-plos-one

## Acknowledgements

We would like to thank the disorientation research group at Institut Jean Nicod for their feedback on some of the material in this article. We would also like to thank Matthew Ratcliffe, Marcella Schmidt di Friedberg, Barbara Tversky and Andreas Falck for their insightful and encouraging feedback.

